# Dissecting Schizophrenia Biology Using Pleiotropy with Cognitive Genomics

**DOI:** 10.1101/2024.04.16.24305885

**Authors:** Upasana Bhattacharyya, Jibin John, Todd Lencz, Max Lam

**Affiliations:** Institute of Behavioral Science, Feinstein Institutes for Medical Research, Manhasset, NY; Division of Psychiatry Research, Zucker Hillside Hospital, Glen Oaks, NY; Departments of Psychiatry and Molecular Medicine, Zucker School of Medicine at Hofstra/Northwell, Hempstead, NY; Institute of Mental Health, Singapore; Lee Kong Chian School of Medicine, Population and Global Health, Nanyang Technological University

**Keywords:** schizophrenia, cognition, educational attainment, GWAS, pleiotropy

## Abstract

Given the increasingly large number of loci discovered by psychiatric GWAS, specification of the key biological pathways underlying these loci has become a priority for the field. We have previously leveraged the pleiotropic genetic relationships between schizophrenia and two cognitive phenotypes (educational attainment and cognitive task performance) to differentiate two subsets of illness-relevant SNPs: (1) those with “concordant” alleles, which are associated with reduced cognitive ability/education and increased schizophrenia risk; and (2) those with “discordant” alleles linked to reduced educational and/or cognitive levels but lower schizophrenia susceptibility. In the present study, we extend our prior work, utilizing larger input GWAS datasets and a more powerful statistical approach to pleiotropic meta-analysis, the Pleiotropic Locus Exploration and Interpretation using Optimal test (PLEIO). Our pleiotropic meta-analysis of schizophrenia and the two cognitive phenotypes revealed 768 significant loci (159 novel). Among these, 347 loci harbored concordant SNPs, 270 encompassed discordant SNPs, and 151 “dual” loci contained concordant and discordant SNPs. Competitive gene-set analysis using MAGMA related concordant SNP loci with neurodevelopmental pathways (e.g., neurogenesis), whereas discordant loci were associated with mature neuronal synaptic functions. These distinctions were also observed in BrainSpan analysis of temporal enrichment patterns across developmental periods, with concordant loci containing more prenatally expressed genes than discordant loci. Dual loci were enriched for genes related to mRNA translation initiation, representing a novel finding in the schizophrenia literature.

---

Schizophrenia is a complex neuropsychiatric disorder affecting approximately 1% of the global population, contributing significantly to the worldwide burden of disease^1^. Over the past decade, genome-wide association studies (GWAS) have identified many genomic loci associated with schizophrenia, revealing its heterogeneous and multifaceted nature^2–6^. However, gleaning biological insights from GWAS results has proven challenging, with downstream analyses pointing broadly toward neuronal and synaptic mechanisms without clear mechanistic differentiation^2,4,7^. Pleiotropic analyses, combining GWAS of schizophrenia with other genetically correlated forms of psychopathology, have boosted the power of gene set enrichment analyses; such studies have implicated a range of neurodevelopmental, synaptic, and other molecular pathways^8,9^ but have not adequately parsed the association of individual pathways with specific phenotypic features.

In two recent studies^10,11^, we have demonstrated that pleiotropic analysis of schizophrenia GWAS with cognitive endophenotypes may permit well-powered, fine-grained, mechanistic differentiation of molecular pathways associated with specific phenotypic constellations. Impaired cognitive ability is a core dimension of schizophrenia pathology^12^; cognitive deficits often emerge early, frequently preceding schizophrenia’s onset by years and contributing to diminished functional outcomes, including educational attainment^13^. Moreover, recent GWAS of cognition and educational attainment demonstrate significant overlap with schizophrenia risk loci^10,14–17^. However, genetic correlation analyses reveal a nuanced relationship between cognitive function, educational attainment, and schizophrenia risk. While there is a significant negative genetic correlation between reduced cognitive function and increased schizophrenia susceptibility (r_g_ ≈ −0.20), there exists a paradoxical, positive genetic correlation between greater educational attainment and schizophrenia risk (r_g_ ≈ 0.10), despite a strong genetic overlap between educational attainment and cognitive ability (r_g_ ≈ 0.70).

Thus, while educational attainment is often viewed as a proxy for cognitive ability, the imperfect genetic concordance between the two can be exploited in pleiotropic analyses of schizophrenia. In our previous work^10,11^, we identified two distinct subsets of schizophrenia-related genetic loci: those with “concordant” alleles that align with expectations—lower cognition/education and increased illness risk—and “discordant” SNPs that show higher educational attainment and/or cognitive ability alongside greater schizophrenia susceptibility. This distinction is obscured when pleiotropy is examined globally, using only genomewide correlational analysis (r_g_). Downstream analyses of concordant loci implicated neurodevelopmental pathways in schizophrenia, while discordant loci were associated with postnatally expressed synaptic mechanisms underlying the disorder.

The present study was designed to replicate and extend our prior pleiotropic meta-analysis, utilizing substantially larger input GWAS for schizophrenia, cognitive task performance, and educational attainment than were available in our prior report^10^. Moreover, in the current investigation, we applied a novel, more powerful meta-analytic approach to pleiotropy, the ‘Pleiotropic Locus Exploration and Interpretation using Optimal test’ (PLEIO)^18^. This methodology permits a direct assessment of the directional associations among these phenotypes. Based on our prior report, we anticipate the identification of novel loci previously unreported in discovery GWAS datasets. Additionally, post-GWAS annotation of these pleiotropically identified loci is expected to extend beyond neurodevelopmental and synaptic pathways, offering deeper insights into the biological mechanisms underpinning these associations (**Figure 1**).

**Figure 1.**
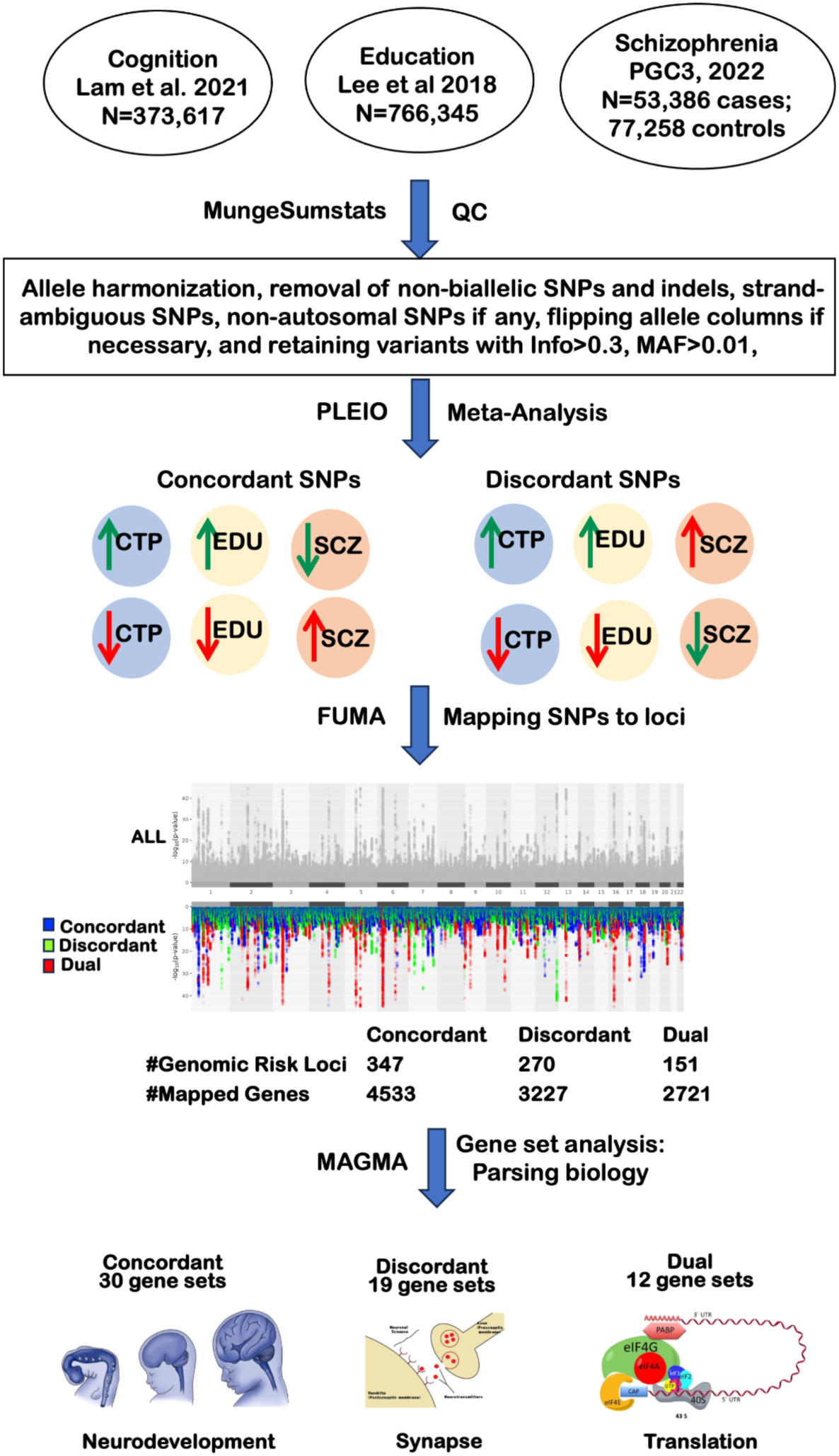
Study strategy and workflow. Methods underlying the present report are broken down into (i) GWAS summary statistics QC phase using ‘mungesumstats,’ (ii) PLEIO pleiotropy meta-analysis, (iii) FUMA GWAS annotation and (iv) MAGMA gene set analysis and further downstream analytic procedures.

Building on previous findings, our current study integrates the latest GWAS on schizophrenia (N= 53,386 cases and 77,258 controls; GWAS mean χ^2^ =2.0126; total SNPs = 7,659,767)^2^, and cognitive function (N=373,617; GWAS mean χ^2^ =2.11; total SNPs = 8,050,310)^11^ to deepen our understanding of its genetic underpinnings. We chose to utilize the educational attainment data set from Lee et al., 2018 (N=766,345; mean GWAS χ²=2.647; total SNPs=10,101,242)^19^, rather than the most recent study of >3M individuals^20^, which could potentially introduce bias into our pleiotropy analyses due to imbalance in sample size and power relative to the schizophrenia and cognitive GWASs. All the GWASs used here are generated from populations of European ancestry.

In the current study, we applied rigorous quality control (QC) to each set of summary statistics utilizing the MungeSumstat tool^21^, which we have optimized as a Python wrapper (accessible via GitHub). We employed the default QC parameters of MungeSumstats^21^, which include removing non-biallelic SNPs and strand-ambiguous SNPs, ensuring consistent direction of reported SNP effects, and ensuring that all variants have non-zero effect sizes and/or standard errors. These QC procedures resulted in the retention of 6,803,445 SNPs common to all three phenotypes, with INFO scores greater than 0.3 and minor allele frequencies above 0.01 (See Supplementary Information).

Two core considerations guided our approach to pleiotropic meta-analysis across the three GWAS summary statistics: (i) We required a method that can account for cryptic sample overlap across GWAS to prevent statistical inflation during meta-analytic procedures; and (ii) we required an approach that could account for SNPs with allelic effects of opposite signs across phenotypes, as our primary aim was to identify “Discordant” as well as “Concordant” SNPs. To our knowledge, only two methods exist that adequately support study objectives: ASSET (Association analysis based on subsets)^22^ and PLEIO (Pleiotropic Locus Exploration and Interpretation using Optimal test)^18^. PLEIO overcomes limitations associated with fixed-effect model approaches by employing a random-effect model to capture genetic correlations and heritabilities across trait pairs^18^. Relative to ASSET, PLEIO is more computationally efficient and provides greater power in simulations across a range of different heritabilities^18^. Linkage disequilibrium score regression (LDSC) implemented within PLEIO estimates genetic covariance and environmental correlation matrices across traits to account for potential sample overlap.

Consistent with our earlier study, we reversed the effect direction per variant for schizophrenia to align with the interpretation of allelic effects on cognition and education^10^; in other words, decreased risk for schizophrenia was coded by positive allelic effects, just as higher scores on cognitive task performance and educational attainment are coded with positive allelic effects. By applying PLEIO, we identified two subsets of variants, concordant and discordant, defined as in our prior study^8^. In the notation that follows, the symbol ∩ indicates variant subsets with the same effect direction across the traits, and the symbol | represents traits whose effect sizes exhibit the opposite direction compared to the other two traits. Thus, the concordant subset includes variants with the same effect direction (i.e., scz ∩ edu ∩ cog, after reversal of sign for schizophrenia). By contrast, the discordant subset includes three distinct sets of variants: (i) edu ∩ cog | scz (schizophrenia outliers, i.e., same allele is associated with an increase in cognitive ability, educational attainment, and schizophrenia risk); (ii) scz ∩ cog | edu (education outliers, i.e., variants associated with an increase in schizophrenia risk and reduced cognitive ability, but increased educational attainment); and (iii) scz ∩ edu | cog (cognition outliers, i.e., variants associated with an increase in schizophrenia risk and reduced educational attainment but increased cognitive ability). Independent genome-wide significant loci (p<5×10^−8^) were identified using the Functional Mapping and Annotation (FUMA) pipeline^23^ (See Data Availability). We excluded the MHC region (chr6:25000000-3500000) from further analysis due to its complex LD patterns, opting to retain a single top SNP from this region.

After harmonization in FUMA (see supplementary methods), we identified three distinct sets of genome-wide significant loci (**Figure 2**): (i) 347 independent loci with exclusively concordant genome-wide significant SNPs (Supplementary Table 1); (ii) 270 independent genomic loci in which all genome-wide significant loci were discordant (Supplementary Table 2); and (iii) 151 “dual” loci, containing both concordant and discordant variants (Supplementary Methods; Supplementary Table 3). The ‘dual’ variant set is novel, ostensibly due to the increased power of the larger input GWAS sample sizes. Next, we compared these results with our prior report^8^, where we reported only Concordant (89 loci) and Discordant (65 loci) with earlier and more modestly sampled versions of the input GWAS.

**Figure 2:**
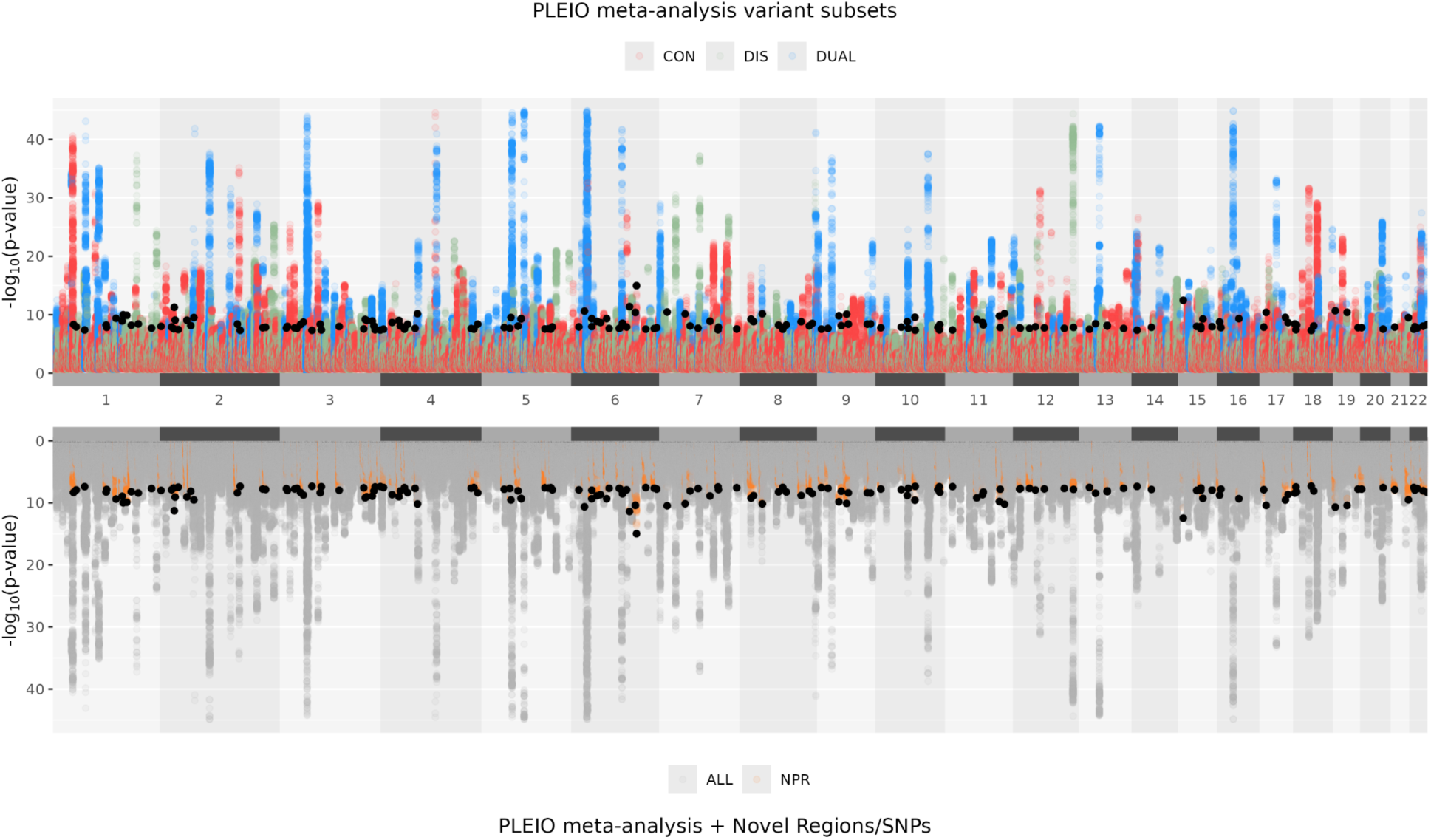
Miami plot of PLEIO meta-analysis results. Top panel: visualization of variant subsets derived from –blup recalibrated effect size directions: (i) Concordant variants (red) (ii) Discordant variants (green) and (iii) Dual variants (blue). Bottom panel: Complete pleiotropic meta-analysis results from the PLEIO tool, NPR: novel loci (orange), Not Previously Reported; Novel index variants reaching genome-wide significance 5×10^−8^ are represented as black dots.

To identify novel loci emerging from our PLEIO meta-analysis, we merged the loci reported for schizophrenia, cognitive task performance, and educational attainment in the original input GWAS publications, along with loci obtained from our pleiotropic meta-analysis using the “merge” option in ‘bedtools’^24^. (We first employed FUMA to map significant variants to loci for education since the original publication lacked comparable locus information.) This process yielded 739 genome-wide significant (p < 5×10^−8^) loci emerging from the pleiotropic meta-analysis, as well as 280 loci for schizophrenia, 432 for education, and 292 for cognitive task performance; these numbers differ slightly from the original reports due to the merging of partially overlapping loci in bedtools (see Supplementary Methods). A Venn diagram of these results is presented in **Figure 3**, revealing that 166 PLEIO loci are not being identified in any of the three input GWASs. By contrast, several loci remained specific to the input GWASs and did not reach genome-wide significance in the pleiotropic meta-analysis (schizophrenia: 94; education: 46; cognitive task performance: 52), presumably indicating that these loci were specific to one phenotype and not pleiotropic.

**Figure 3:**
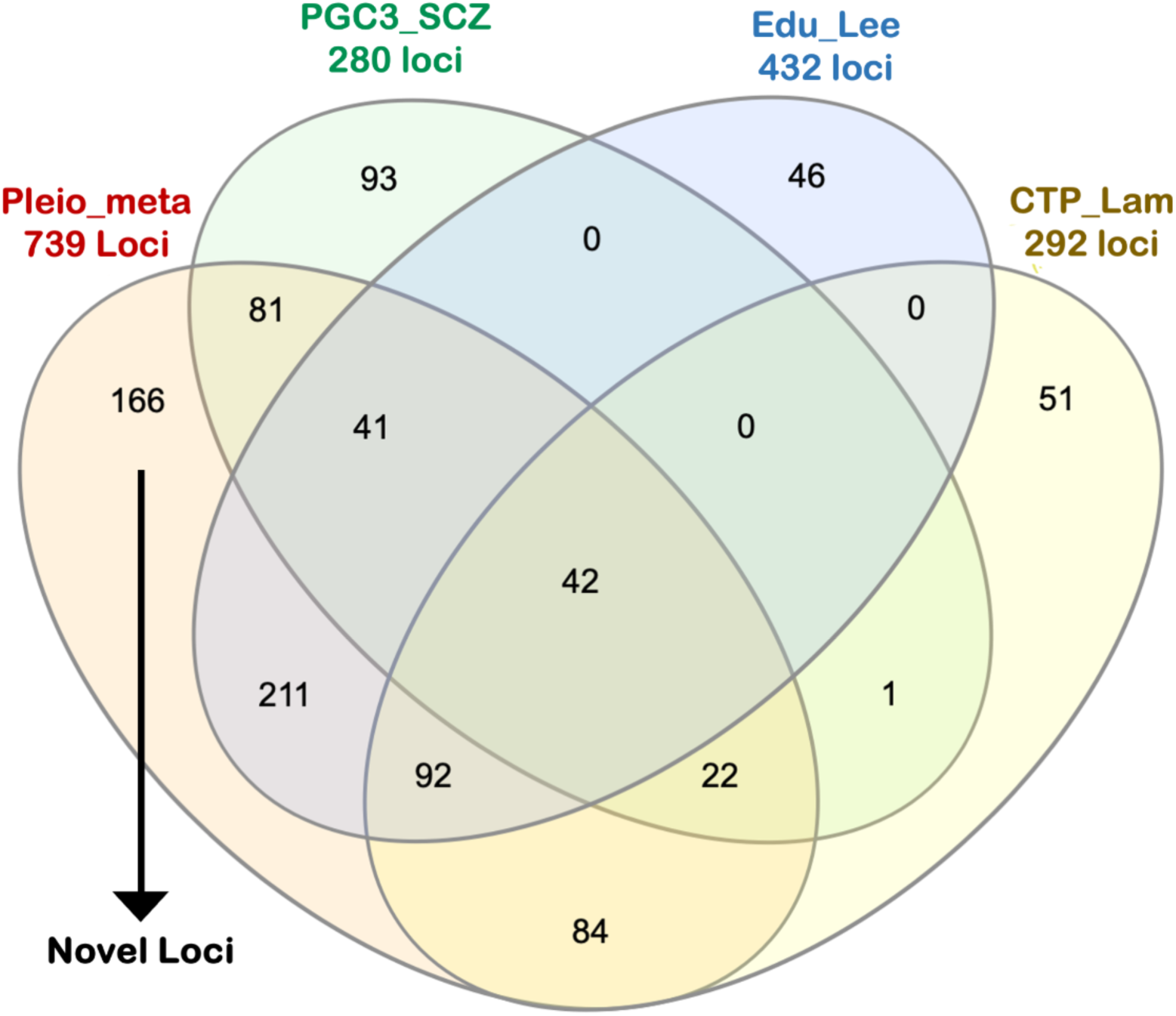
Venn Diagram Comparing Significant PLEIO Loci to Significant Loci from Input GWASs Pleio_meta: results from PLEIO meta-analysis; SCZ_PGC3: Schizophrenia GWAS input (PGC Schizophrenia Working Group^2^); Edu_Lee: Education GWAS^19^; CTP_Lam: Cognitive task performance GWAS^11^.

Comparing results with our earlier pleiotropic meta-analytic study^8^, we identified 110 novel loci. Notably, 89 of these loci (comprising 80.9%) were validated as significant in subsequent, larger GWAS datasets for the analyzed traits, underscoring the efficacy of pleiotropic analysis in uncovering novel genetic loci^2,11,19^. In the present report, we employed the most recent GWAS data for schizophrenia and cognitive task performance, leaving us without newer studies for direct comparisons. Nevertheless, we can cross-reference a more recent study on educational attainment^20^ with sample sizes nearly three times larger than our input GWAS^19^. Because the complete summary statistics of the recent education GWAS are not available in the public domain, we have used the restricted summary statistics for the complete additive autosomal GWAS that contains clumped results for 8,618 variants with P < 1×10^−5^ (Sample size = 3,037,499). To conduct the look-up, we checked if any loci identified from our analysis overlap with 20kb upstream or downstream of any reported variants. Among the 166 novel loci identified, 111 (66.86%) were confirmed in the latest educational GWAS, validating the PLEIO methodology’s effectiveness and highlighting its increased capacity for discovering novel genetic loci across phenotypes (Supplementary Table 4). It is important to note that we exclusively considered variants achieving genome-wide significance (adjusted P < 1.28×10^−8^) to ensure compatibility with our dataset.

MAGMA (version 1.10) (Multi-marker Analysis of GenoMic Annotation)^25^ gene set analysis was conducted to identify specific biological processes underpinning Concordant, Discordant and ‘Dual’ variant sets. Variants were annotated to gene regions, setting boundaries 35 kb upstream and 10 kb downstream to include likely regulatory regions associated with each gene, as suggested by previous publications^4,26–28^. Following annotation, gene-based genome-wide association (MAGMA-GBGWA) analysis was conducted using the SNP-wise mean method to compute the p-value for each gene. The gene-based results were subsequently entered into the MAGMA competitive gene set analysis. Gene sets from MsigDb C5 collection (GO-basic obo file released on 2023-07-27) that contain all the Gene Ontology gene sets^29^ (GO Biological Process ontology, GO Cellular Component ontology, GO Molecular Function ontology) were utilized as annotations for gene set analysis. We excluded genes within chromosome X or Y or extended MHC region. Any gene set that contained less than 10 genes was also excluded (See Supplementary Methods). Competitive gene set analyses were performed separately for Concordant, Discordant, and Dual variants against 7,353 gene sets (See Data Availability).

MAGMA competitive gene set analysis for the concordant loci revealed 30 GO annotations enriched at FDR<0.05 (Supplementary Table 5). Results were broadly consistent with our previous observation that concordant loci were enriched for transcriptional regulation pathways and neurodevelopmental genes^8^. With the current study’s enhanced power, we could identify multiple RNA regulatory pathways implicated at concordant loci beyond the chromatin regulation discussed in our prior report. These include a broader range of gene sets involved with chromosome organization, chromatin formation, and methylation. Additionally, we identified significant gene sets regulating the spliceosome, which has emerged as a critical focus of abnormalities in neuropsychiatric disorders^30^.

The present results also provided additional details on the nature of neurodevelopmental processes implicated by concordant genes, including macro-level processes such as head development as well as cellular processes of neurogenesis. Notably, two FDR-significant gene sets specifically implicated forebrain development. As the forebrain develops into the cerebral cortex, neurodevelopmental models of schizophrenia have long focused on this process^31^. Converging lines of evidence demonstrate that illness-related cognitive impairment in schizophrenia is neurodevelopmental in origin and long predates overt symptomatology^32,33^. Forebrain developmental abnormalities may provide a genetic basis for the increased rate of craniofacial abnormalities in patients with schizophrenia, which are sometimes also observed in their first-degree relatives^34^. Indeed, it has been suggested that refined measurement of craniofacial dysmorphology may represent a readily accessible index of neurodevelopmental abnormalities in schizophrenia^35,36^. Additionally, very recent studies have recapitulated gene expression abnormalities in schizophrenia using forebrain organoids derived from induced pluripotent stem cells of patients with schizophrenia^34^.

MAGMA competitive gene set analysis at discordant loci revealed 19 GO annotations enriched at FDR<0.05 (Supplementary Table 5). As in our prior study^8^, the discordant subset demonstrated significant enrichment in synaptic pathways, especially the postsynaptic density, which has emerged as perhaps the strongest gene set emerging from schizophrenia GWAS^2^. The present results suggest that these pathways are distinct from those causing the cognitive deficits related to schizophrenia and are instead associated with the counter-intuitive positive genetic correlation of schizophrenia with educational attainment. As noted in our prior report^8^, these results do not point towards a subset of schizophrenia patients marked by high educational attainment but rather may indicate an inverted-U function: post-synaptic processes that underlie successful academic performance may also lead to illness if taken too far.

With the enhanced power of the present study, we identified several novel gene sets enriched in the discordant loci, most notably several involved in neurodevelopment specific to the hindbrain/cerebellum. While hindbrain development has not been a primary focus in schizophrenia research, abnormalities in the mature cerebellum have been identified using both structural and functional MRI^37^. Traditionally linked to motor control, the cerebellum is increasingly recognized for its involvement in cognitive and emotional functions^38^. Identification of cerebellar abnormalities, in conjunction with synaptic deficits, suggests a previously under-appreciated role for metabotropic^39^ and delta ionotropic^40^ glutamate receptors in the illness process.

Several additional gene sets were enriched in discordant loci, pointing towards novel targets for future study. The regulation of the Notch signaling pathway plays a pivotal role in governing cell fate decisions during neurodevelopment but is increasingly understood as subserving learning processes in mature neurons^41^. The unfolded protein response pathway maintains homeostasis within the endoplasmic reticulum (ER), in response to ER stress^42^; molecular evidence of dysfunction in this system has recently been observed in postmortem brains of patients with schizophrenia^43^.

In gene set enrichment analysis for “dual” loci, pathways related to translation initiation were the primary significant result (Supplementary Table 5). Intriguingly, one of the genes driving this association was *EIF3C*, which neighbors (and is often included in) the schizophrenia-associated copy number variant at distal 16p11.2^44^. The translation initiation pathway is fundamental to many biological processes, as the effective operation of diverse cell types hinges on the precise and dynamic regulation of mRNA translation to proteins. Neurons, characterized by their highly polarized morphology, particularly depend on the spatial organization of mRNA translation machinery^45^. Moreover, temporal control of mRNA translation is pivotal for neurons responding promptly to environmental changes by modifying synaptic protein composition; this process is essential to experience-dependent long-term synaptic plasticity^46^. A recent study directly investigated the translational control of protein synthesis in olfactory neurosphere-derived (ONS) cells drawn from patients with schizophrenia and controls. The study found 48 differentially expressed eIF2 pathway proteins and mRNA transcripts associated with schizophrenia; additionally, mTOR and eIF4 signaling pathways, known regulators of protein synthesis, were implicated^47^. Thus, our finding that translation-related gene sets are enriched at the “dual” loci highlights the need for more studies to understand the role of protein translation in schizophrenia.

Finally, we utilized the publicly accessible BrainSpan dataset^48^ to analyze temporal gene expression patterns concerning the three sets of pleiotropic loci. We hypothesized that genes at concordant loci, primarily involved in neurodevelopment, would peak in expression prenatally. In contrast, genes linked to adult synaptic regulation (discordant genes) would be more active during later developmental stages. We transformed neurodevelopmental stages into weeks to uncover more intricate gene expression profiles over time. As hypothesized, we found substantial time by locus-type interaction effects among gene sets, with discordant genes demonstrating an upward slope over time relative to concordant genes (Concordant vs Discordant, b = 1.92×10^−4^, s.e. 4.79×10^−5^, p = 6.41×10^−5^) (**Figure 4**). Somewhat surprisingly, genes at the dual loci demonstrated an even greater downward slope relative even to concordant genes (Concordant vs Dual, b = 2.45×10^−4^, s.e. = 4.76×10^−4^, p = 3.14×10^−7^; Discordant v Dual, b = 4.37×10^−4^, s.e. = 5.47×10^−5^, p = 3.63×10^−15^). However, the main effect for developmental weeks was not statistically significant for any of the three sets of genes individually (Concordant, b = –6.49×10^−5^, s.e. = 2.5×10^−4^ p = 0.796; Discordant, b = 0.000138, s.e. = 0.000243, p = 0.572; Dual, b = –0.000375, s.e.= 0.000238, p = 0.124).

**Figure 4.**
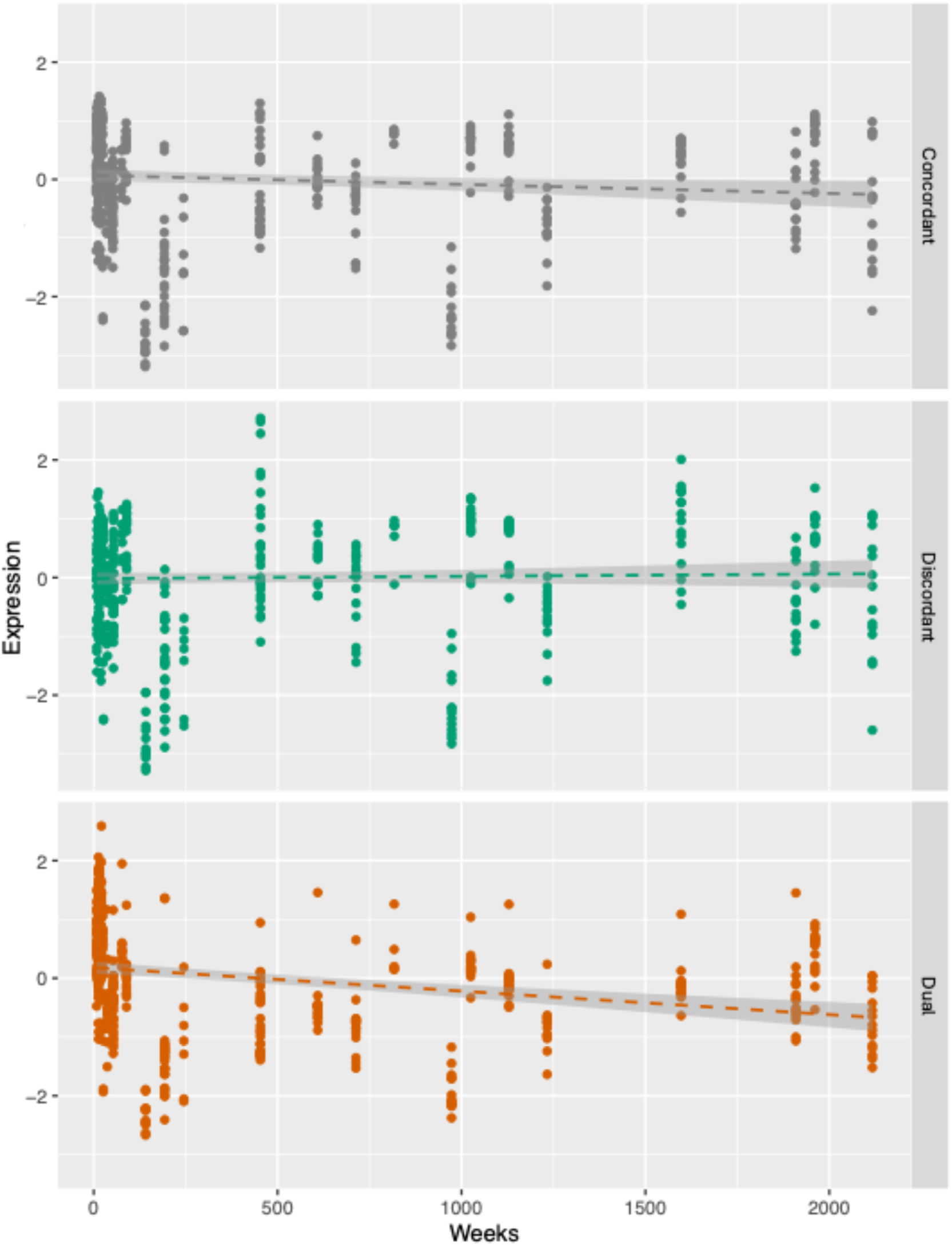
Brainspan Temporal Gene Expression Profiles. Top panel: Brainspan gene expression profiles for ‘Concordant’ genes. Middle panel: Brainspan gene expression profiles for ‘Discordant’ genes. Bottom panel: Brainspan gene expression profiles for ‘Dual’ genes. x-axis: Development time in Weeks. y-axis: standardized (z-scores) of normalized gene expression values in BrainSpan.

In conclusion, our investigation has yielded several key findings that replicate and extend our prior report^8^ on the pleiotropic relationship between schizophrenia, cognitive task performance, and educational attainment. First, applying pleiotropic analysis enabled us to harness the collective power of multiple GWAS datasets for enhanced locus discovery. Second, our findings reaffirm the divergence between forebrain neurodevelopmental pathways and synaptic regulation in schizophrenia. While these pathways have previously emerged as the primary results from gene set analysis of common^4^ and rare^49^ genetic variation in schizophrenia, our work indicates that these can be parsed with respect to their endophenotypic manifestations. The larger sample sizes in the current report permit us to expand and refine these pathways, newly demonstrating roles for altered gene splicing at concordant loci and notch signaling at discordant loci. Third, identifying newly emergent gene set categories related to cerebellar development at discordant loci and translational initiation factors at dual loci provides clues to schizophrenia pathophysiology and potential treatment targets not previously reported in genetics literature.

Our pleiotropic integration of psychiatric GWAS with endophenotypic GWAS was designed to parse biological mechanisms underlying schizophrenia. While our primary results contribute to our understanding of the etiopathophysiology of the disorder, pointing towards novel treatment targets, we propose that the resulting data may, in the future, have potential clinical applicability in the form of more refined polygenic risk scores (PRS) reflecting these distinct biological pathways. Such an approach has been successfully developed in recent studies of Type 2 diabetes (T2D), in which the genetic heterogeneity of that complex disease has been parsed into several dissociable biological processes (e.g., lipodystrophy, insulin synthesis, hepatic lipid metabolism, etc.)^50^. The model, as proposed by Udler^51^ and McCarthy^52^, is not that there are multiple distinct subtypes of T2D, but rather that the observed diagnostic entity is comprised of several underlying dimensions and that any individual diagnosed with T2D has some combination of risk along these dimensions. Each dimension of risk is quantifiable by subsets of SNPs drawn from the overall PRS for T2D, corresponding to genetic regions relevant to the underlying biological processes, and a patient’s PRS profile along these various dimensions can more accurately differentiate outcomes in T2D^50^. Recent endophenotype studies in schizophrenia have been guided by a categorical approach to the identification of ‘biotypes,’ such as a subtype of psychosis marked by cognitive deficits^53^ and distinct brain dysmorphism^54^. While the present results are consistent with the evidence of such endophenotypic manifestations in some individuals with schizophrenia, our approach suggests that these represent dimensions of abnormality rather than distinct subgroups of patients.

## Data and Code Availability

1. GWAS summary statistics for pleiotropy analysis and gene set analysis

a. GWAS: gs://pleiotropy-proj-1/02-interim-testing/MTAG_PLEIO/PLEIO_EDU_Cog_SCZ_ldsc_vcf/PLEIO_Output
b. FUMA: gs://pleiotropy-proj-1/02-interim-testing/Post_GWAS_Anlalysis/FUMA_results/PLEIO.PLEIO_FUMA_job240552
c. MAGMA: gs://pleiotropy-proj-1/NoMHC_Extended_25Mb_35Mb
d. Summary statistics and other auxiliary files to be made available upon acceptance of the manuscript
2. Github links

a. https://github.com/mlamcogent/cogent-data-curation/tree/main
3. Publicly available GWAS summary statistics

a. SSGAC https://thessgac.com
b. Cognitive Task Performance (Lam et al., 2022) https://storage.googleapis.com/broad_institute_mlam/brainstorm-v2-local-gencor-1/03_quality_control_sumstatsqc/07_Data_Release_GWAS_Catalog_01/Lam_et_al_2021_CognitiveTaskPerformance.tsv.gz
c. PGC3 https://pgc.unc.edu/for-researchers/download-results/
4. Bioinformatic Tools & Resources

a. Bedtools https://github.com/arq5x/bedtools2?tab=readme-ov-file
b. Mungesumstats https://github.com/neurogenomics/MungeSumstats
c. PLEIO https://github.com/cuelee/pleio
d. MSigDB https://www.gsea-msigdb.org/gsea/msigdb/
e. BrainSpan https://www.brainspan.org/

## Supporting information

supplementary section

## Data Availability

Data and Code Availability
1. GWAS summary statistics for pleiotropy analysis and gene set analysis
a. GWAS: gs://pleiotropy-proj-1/02-interim-testing/MTAG_PLEIO/PLEIO_EDU_Cog_SCZ_ldsc_vcf/PLEIO_Output
b. FUMA: gs://pleiotropy-proj-1/02-interim-testing/Post_GWAS_Anlalysis/FUMA_results/PLEIO.PLEIO_FUMA_job240552
c. MAGMA: gs://pleiotropy-proj-1/NoMHC_Extended_25Mb_35Mb
d. Summary statistics and other auxiliary files to be made available upon acceptance of the manuscript
2. Github links
a. https://github.com/mlamcogent/cogent-data-curation/tree/main
3. Publicly available GWAS summary statistics
a. SSGAC https://thessgac.com
b. Cognitive Task Performance (Lam et al., 2022) https://storage.googleapis.com/broad_institute_mlam/brainstorm-v2-local-gencor-1/03_quality_control_sumstatsqc/07_Data_Release_GWAS_Catalog_01/Lam_et_al_2021_CognitiveTaskPerformance.tsv.gz
c. PGC3 https://pgc.unc.edu/for-researchers/download-results/
4. Bioinformatic Tools & Resources
a. Bedtools https://github.com/arq5x/bedtools2?tab=readme-ov-file
b. Mungesumstats https://github.com/neurogenomics/MungeSumstats
c. PLEIO https://github.com/cuelee/pleio
d. MSigDB https://www.gsea-msigdb.org/gsea/msigdb/
e. BrainSpan https://www.brainspan.org/

## Acknowledgments

This work was supported by the National Institute of Mental Health of the National Institutes of Health (NIH) under award no. R01MH117646 (T.L., principal investigator). The content is solely the responsibility of the authors and does not necessarily represent the official views of the NIH.

