## supplementary section for "Dissecting Schizophrenia Biology Using Pleiotropy with Cognitive Genomics"

Supplementary Methods

The subsequent sections detail our methodological approach, encompassing data curation, pleiotropic meta-analysis, identification of independent genome-wide significant loci, labeling of loci in terms of the presence of concordant and discordant variants, and characterization of meta-analytic loci relative to input GWAS. Additionally, we outline our gene set analysis technique and examine these gene sets using temporal gene expression profiles in the brain.

### 1. Data Curation

We initiated the data curation process by collecting the most recent genome-wide association study (GWAS) summary statistics for the relevant traits. All GWAS summary statistics used in this study consisted of individuals with European ancestry^1–3^. Specifically, for schizophrenia, we obtained the GWAS summary statistics from the Psychiatric Genomics Consortium (<https://pgc.unc.edu/for-researchers/download-results/>; N= 53,386 cases and 77,258 controls; GWAS mean χ2 =2.0126; total Variants = 7,659,767)^3^. The GWAS summary statistics for educational attainment were acquired from the Social Science Genomics Association Consortium (SSGAC;https://thessgac.com) (N=766,345; mean GWAS χ²=2.647; total Variants=10,101,242)^2^. This GWAS was selected, rather than the most recent educational attainment GWAS^4^, to maintain a sample size comparable to the other GWASs included in the current report. We utilized the summary statistics from our previous meta-analytic publication for cognitive task performance, the largest available for the phenotype (N=373,617; GWAS mean χ2 =2.11; Total Variants = 8,050,310)^1^.

We implemented rigorous quality control (QC) protocols on the GWAS summary statistics using the MungeSumstat tool^5^. This process entailed a thorough suite of QC checks designed to rectify common genetic data inconsistencies, including allele discrepancies, strand ambiguity, and anomalies in chromosome nomenclature. We further enhanced the uniformity of GWAS summary statistics by standardizing strand designation, SNP ID, reference genome build, effect allele, p-value, and effect size formats. We ensured every variant displayed a nonzero effect size and/or standard error, with sample counts (N) maintained as integers across all datasets. Our QC measures also eliminated non-biallelic SNPs, indels, and SNPs with INFO scores below 0.3 or minor allele frequencies under 0.001. Following these rigorous quality control steps, we retained 6,803,445 SNPs consistent across the three phenotypes and available for subsequent analysis.

### 2. Meta-Analysis

We then utilized PLEIO to conduct a cross-phenotype pleiotropic meta-analysis to identify shared genetic loci between schizophrenia, cognition, and education. PLEIO has been demonstrated^6^ to have several advantages over the ASSET^7^ (Association analysis based on subsets) method, which we used in our prior report^8^. PLEIO overcomes limitations associated with fixed-effect model approaches by employing a random-effect model that captures genetic correlations and heritabilities across trait pairs, maximizing statistical power^6^. Additionally, PLEIO enhances computational efficiency relative to other approaches by employing spectral decomposition on the covariance matrix. PLEIO utilizes a variance component test to identify pleiotropic SNPs that influence multiple traits simultaneously, incorporating an importance sampling method to provide an accurate p-value estimate.

In this analysis, we have utilized the *ldsc_preprocess.py* script implemented in PLEIO that calls the LDSC software^6,8^ (<https://github.com/bulik/ldsc>) to estimate both genetic covariance and environmental correlation matrices across traits - at the same time generating GWAS summary statistics with the standardized effect sizes, before carrying out the pleiotropic meta-analysis. We then used the *Pleio.py* script (<https://github.com/cuelee/pleio>) implemented in PLEIO to perform the pleiotropic meta-analysis. Specific codes and commands are provided in the GitHub repository (https://github.com/mlamcogent/cogent-pleiotropy-proj/tree/main)

### 3. Identification of Subsets of Variants with Homogenous Relationships across Schizophrenia, Educational Attainment, and Cognitive Ability

PLEIO employs a recalibration process to calculate trait-specific beta values and estimate their standard errors using the *--blup* option during the meta-analysis^6^. Consistent with our earlier report^9^, we utilized the directionality of the recalibrated beta effect sizes generated as part of the pleiotropic meta-analytic procedures to identify subsets of variants genome-wide across schizophrenia, cognitive task performance, and educational attainment. As with our earlier report, the direction of effect sizes for each variant was reversed within the schizophrenia GWAS summary statistics to align with the interpretation of cognition and education scores, such that a negative allele effect would indicate risk for schizophrenia and a positive allele effect would suggest a protective effect to schizophrenia.

Variants were then organized into subsets described in detail in our prior report^9^. Also consistent with our earlier report^9^, the symbol ∩ designates a variant with the same effect direction across any two phenotypes (after the reversal of schizophrenia). In contrast, the symbol | denotes variants in which one phenotype displays an effect direction that opposes those of the other two traits (again, after the reversal of schizophrenia). Variants with beta close to zero (between 0.001 and -0.001) for a given input trait were considered to have no direction of effect for that trait. Thus, we define one subgroup of variants as "Concordant" if allelic effects associated with increased cognitive ability and educational attainment are associated with a decrease in schizophrenia risk (scz ∩ edu ∩ cog). By contrast, "Discordant" variants, as in our prior report, are comprised of three subsets: (1) edu ∩ cog | scz (schizophrenia outliers, which have alleles associated with an increase in cognitive ability, educational attainment, and schizophrenia risk); (2) scz ∩ cog | edu (education outliers, where alleles associated with an increase in schizophrenia risk and reduced cognitive ability are also associated with increased educational attainment) and (3) scz ∩ edu | cog (cognition outliers, with alleles associated with an increase in schizophrenia risk and reduced educational attainment, but also increased cognitive performance). These subgroups are further represented visually in Supplementary Fig 1.

### 4. Identification and Consolidation of Independent Loci

Independent genome-wide-significant loci for each PLEIO meta-analysis subset (Concordant and DIscordant) were identified using the FUnctional Mapping and Annotation (FUMA; version 1.5.2) pipeline. We have used the default parameters of FUMA to perform double clumping of SNPs based on the European 1000 Genomes Project Phase 3 reference panels. The clumping of variants to identify significant genome-wide loci was done in two steps. First, the clumping parameter was set to clump SNPs with *p*-value < 5x10^‐8^ and an LD of r^2^ < 0.6 to identify “independent significant SNPs.” The second clumping step identified ‘lead SNPs” using r^2^ < 0.1 to clump independent significant SNPs with the strongest signal at a given locus. The MHC region (Chr6:25MB-35MB) was excluded from the clumping analysis; due to the highly complex LD structure within the region, only a single top SNP from this region was retained. We included SNPs from the 1000 Genomes reference panel represented within the given locus to maximize the potential for biological annotation within identified loci. LD blocks of independent significant variants 250 kb or less were merged into a single genomic locus. We identified 503 loci harboring concordant SNPs and 428 loci discordant SNPs. However, some overlap existed between these categories; following the procedure followed in our prior report^10^, we harmonized the loci obtained from the two subsets using default settings of the “*merge*” function implemented in the “bedtools” package (Version 2; <https://github.com/arq5x/bedtools2>)^10^ i.e, merging loci overlapping each other by at least 1 base pair. While harmonizing across significant genomic loci, we found that many contained both concordant and discordant variant subsets. For the current report, these are called "Dual"loci. For subsequent analysis, we subgrouped the pleiotropic meta-analysis summary statistics from PLEIO into three distinct groups of SNPs: (i) those within concordant loci, (ii) those solely within discordant loci, and (iii) those present in the dual loci.

### 5. Validation of Novel Loci

Consistent with our earlier work^9^, we sought to validate the novel loci against more recent summary statistics to demonstrate that an outcome of investigating pleiotropy is increased discovery power. As we are presently already using the latest and most well-powered GWAS summary statistics input for schizophrenia and cognitive task performance, it would be out of the scope of the current report to validate novel loci in those traits. Nonetheless, we were able to check if any of the novel loci appeared in the recent educational attainment GWAS^20^, which has almost three times the sample size of the current one^2^. However, in the recent educational GWAS, full locus data (including all SNPs at each significant locus) were not reported, and complete summary statistics, including 23andMe, were unavailable. Thus we downloaded the file EA4_additive_p1e-5_clumped.txt from <https://www.thessgac.org/>, which contains additive GWAS meta-analysis of all discovery cohorts for approximately 8,618 independent (pairwise r^2^ < 0.1) autosomal variants with P<1x10^-5^ and N>500,000 (total sample size = 3,037,499). To conduct the look-up, we selected genome-wide significant variants (adjusted P<1.29x10^-8^). We checked if any loci identified from our analysis overlap within 20kb upstream or downstream of any reported SNPs.

### 6. MAGMA gene set analysis

Gene set analyses were performed for each of the three subsets (Concordant, Discordant, and Dual) separately using MAGMA v1.10, consisting of three steps: annotation, to map SNPs onto genes; gene analysis, to compute gene p-values; and finally, gene set analysis. We utilized the gene location file (build 37) of protein-coding genes obtained from https://ctg.cncr.nl/software/magma for annotation of SNPs to genes in MAGMA. We defined the boundaries of the genes as 35 kb upstream and 10 kb downstream to include likely regulatory regions associated with these genes^11–14^ Following annotation, gene-level analysis was conducted, and a snp-wise mean model was used to compute the p-value for each gene. The gene sets were obtained from the most recent GO annotations presented in the GO-basic obo file released on 2023-07-27 and NCBI gene2go annotations downloaded on 2023-09-01 by MSigdb. We used the C5 collection of MSigdb (MSigDB v2023.1.Hs). These annotations are based on specific references that describe the experiments or analyses supporting the association between a particular gene product and a corresponding Gene Ontology (GO) term, along with an evidence code to indicate the level of support for the association with the given GO term. In this subcollection, the gene sets are identified with prefixes such as "GOBP" (Biological Process), "GOMF" (Molecular Function), or "GOCC" (Cellular Component), indicating their respective sources in the Gene Ontology. We removed any genes belonging to chromosome X or Y or extended MHC region (Chr6:25000000-35000000), as these regions were not included in the meta-analysis. We then removed any gene sets that contained less than 10 genes. Finally, MAGMA competitive gene set analysis was conducted against 7,353 gene sets. Due to considerable redundancy across GO annotations, a strict Bonferroni correction would be overly conservative; consequently, we employed a false discovery rate threshold (FDR<0.05) using the Benjamini-Hochberg procedure^15^.

**7. Follow-up Temporal Gene Expression Analysis via BrainSpan**

Follow-up annotation of gene set enrichment results was conducted via publicly available BrainSpan data (<https://www.brainspan.org/static/download.html>). Detailed methodology describing the pre-processing and curation of the BrainSpan data has been described elsewhere^16,17^. MAGMA gene set enrichment outputs were post-processed by selecting only Gene Ontology (GO) gene sets that showed significant association at FDR < 0.05 separately for Concordant, Discordant, and Dual loci. Gene-based p-values for each gene list were vectorized and converted to -logP values for subsequent weighting of expression values. We recoded the developmental stage into “Weeks'' of development to obtain a higher spatial-temporal gene expression profile resolution, as we have done previously^1^. We aggregated the gene expression for a given gene list across brain regions to estimate mean gene expression over time, yielding a whole-brain gene expression estimate. In addition, gene expression for each gene list was weighted by the gene-based -log10 P-values and then normalized by the total gene expression across the BrainSpan data set. Final gene expression profiles were then expressed as gene expression Z-scores. To examine differences in gene expression for each Concordant, Discordant, and Dual gene lists, we carried out linear mixed model analysis for each pair of expression profiles over time (i) Concordant-Discordant (ii) Dual-Discordant (iii) Dual-Concordant. Based on our expectations, the gene expression profile for the Concordant genes, with neurodevelopmental enrichment, would follow a negative gradient; conversely, the gene expression profile for Discordant genes would be follow a positive gradient. To evaluate the hypothesis, we built an interaction model with trait pair (Concordant v Discordant; Dual v Concordant; Dual v Concordant) * Time. We report both the main effect and interaction effects for the trait pairs indicated.

### Supplementary Table Captions

**Supplementary Table 1a**: GWAS significant Pleiotropic Genomic Risk Loci for Concordant Variants

*Note:* Genomic locus : Index of genomic rick loci; uniqID : Unique ID of SNPs consisting of chr:position:allele1:allele2 where alleles are alphabetically ordered; rsID : rsID of the top lead SNP based on dbSNP build 146; chr : chromosome of top lead SNP; pos : position of top lead SNP on hg19; P-value : P-value of top lead SNP (from the input file); start : Start position of the locus; end : End position of the locus; nSNPs : The number of unique candidate SNPs in the genomic locus, including non-GWAS-tagged SNPs (which are available in the user selected reference panel). Candidate SNPs are all SNPs that are in LD (give user-defined r2) with any of independent significant SNPs and either have a P-value below the user defined threshold or are only available in 1000G; nGWASSNPs : The number of unique GWAS-tagged candidate SNPs in the genomic locus which is available in the GWAS summary statistics input file. This is a subset of "nSNPs"; nIndSigSNPs : The number of the independent (at user defined r2) significant SNPs in the genomic locus; IndSigSNPs : rsID of the independent significant SNPs in the genomic locus; nLeadSNPs : The number of lead SNPs in the genomic locus. Lead SNPs are subset of independent significant SNPs at r2 0.1; LeadSNPs : rsID of lead SNPs in the genomic locus.

**Supplementary Table 1b**: Annotations for variants within GWAS significant Pleiotropic Genomic Risk Loci for Concordant Variants

*Note:* uniqID : Unique ID of SNPs consisting of chr:position:allele1:allele2 where alleles are alphabetically ordered; rsID : rsID based on dbSNP build 146; chr : chromosome; pos : position on hg19; effect_allele : Effect/risk allele if it is provided in the input GWAS summary statistics file. If not, this is the alternative (minor) allele in 1000G; non_effect_allele : Non-effect/non-risk allele if it is provided in the input GWAS summary statistics file. If not, this is the reference (major) allele in 1000G; MAF : Minor allele frequency computed based on 1000G; gwasP : P-value provided in the input GWAS summary statistics file. Non-GWAS tagged SNPs (which do not exist in input file but are extracted from the reference panel) have "NA" instead; or : Odds ratio provided in the input GWAS summary statistics file if available. Non-GWAS tagged SNPs (which do not exist in input file but are extracted from the reference panel) have "NA" instead; beta : Beta provided in the input GWAS summary statistics file if available. Non-GWAS tagged SNPs (which do not exist in input file but are extracted from the reference panel) have "NA" instead; se : Standard error provided in the input GWAS summary statistics file if available. Non-GWAS tagged SNPs (which do not exist in input file but are extracted from the reference panel) have "NA" instead; r2 : The maximum r2 of the SNP with one of the independent significant SNPs; IndSigSNP : rsID of the independent significant SNP which has the maximum r2 with the SNP; Genomic locus : Index of the genomic risk loci matching with "Genomic risk loci" table; Novel : Whether the index SNP/locus is novel to the pleiotropic analysis, and if the variant/locus has been validated in recent education attainment GWAS (information from Supplementary Table 4); nearestGene : The nearest Gene of the SNP based on ANNOVAR annotations. Note that ANNOVAR annotates "consequence" function by prioritizing the most deleterious annotation for SNPs which are locating a genomic region where multiple genes are obverlapped. Genes are ecoded in symbol, if it is available otherwise; Ensembl ID. Genes include all transcripts from Ensembl gene build 85 including non-protein coding genes and RNAs; dist : Distance to the nearest gene. SNPs which are locating in the gene body or 1kb up- or down-stream of TSS or TES have 0; func : Functional consequence of the SNP on the gene obtained from ANNOVAR. For exonic SNPs, detailed annotation (e.g. non-synonymous, stop gain and so on) is available in the ANNOVAR table (annov.txt); CADD : CADD score which is computed based on 63 annotations. The higher the score, the more deleterious the SNP is. 12.37 is the suggested threshold by Kicher et al (2014); RDB : RegulomeDB score which is a categorical score (from 1a to 7). 1a is the highest score for SNPs with the most biological evidence to be a regulatory element; minChrState : The minimum 15-core chromatin state across 127 tissue/cell type; commonChrState : The most common 15-core chromatin state across 127 tissue/cell types; posMapFilt : Whether the SNP was used for eQTL mapping or not. 1 is used, otherwise 0. When eqtl mapping is not performed, all SNPs have 0.

**Supplementary Table 1c:** GWAS significant variants within r^2^ = 0.6 of Concordant sentinel variants

*Note:* No : Index of independent significant SNPs; Genomic Locus : Index of assigned genomic locus matched with "Genomic risk loci" table. Multiple independent lead SNPs can be assigned to the same genomic locus; uniqID : Unique ID of SNPs consisting of chr:position:allele1:allele2 where alleles are alphabetically ordered; rsID : rsID based on dbSNP build 146; chr : chromosome; pos : position on hg19; P-value : P-value (from the input file); nSNPs : The number of SNPs in LD with the lead SNP given r2, including non-GWAS-tagged SNPs (which are extracted from 1000G); nGWASSNPs : The number of GWAS-tagged SNPs in LD with the lead SNP given r2. This is a subset of "nSNPs".

**Supplementary Table 1d**: GWAS significant Pleiotropic Concordant sentinel variants within a given genomic locus

*Note:* No : Index of lead SNPs; Genomic Locus : Index of assigned genomic locus matched with "Genomic risk loci" table. Multiple lead SNPs can be assigned to the same genomic locus; uniqID : Unique ID of SNPs consisting of chr:position:allele1:allele2 where alleles are alphabetically ordered; rsID : rsID based on dbSNP build 146; chr : chromosome; pos : position on hg19; P-value : P-value (from the input file); nIndSigSNPs : Number of independent significant SNPs which are in LD with the lead SNP at r2 0.1; IndSigSNPs : rsID of independent significant SNPs which are in LD with the lead SNP at r2 0.1.

**Supplementary Table 2a**: GWAS significant Pleiotropic Genomic Risk Loci for Discordant Variants

*Note:* Genomic locus : Index of genomic rick loci; uniqID : Unique ID of SNPs consisting of chr:position:allele1:allele2 where alleles are alphabetically ordered; rsID : rsID of the top lead SNP based on dbSNP build 146; chr : chromosome of top lead SNP; pos : position of top lead SNP on hg19; P-value : P-value of top lead SNP (from the input file); start : Start position of the locus; end : End position of the locus; nSNPs : The number of unique candidate SNPs in the genomic locus, including non-GWAS-tagged SNPs (which are available in the user selected reference panel). Candidate SNPs are all SNPs that are in LD (give user-defined r2) with any of independent significant SNPs and either have a P-value below the user defined threshold or are only available in 1000G; nGWASSNPs : The number of unique GWAS-tagged candidate SNPs in the genomic locus which is available in the GWAS summary statistics input file. This is a subset of "nSNPs"; nIndSigSNPs : The number of the independent (at user defined r2) significant SNPs in the genomic locus; IndSigSNPs : rsID of the independent significant SNPs in the genomic locus; nLeadSNPs : The number of lead SNPs in the genomic locus. Lead SNPs are subset of independent significant SNPs at r2 0.1; LeadSNPs : rsID of lead SNPs in the genomic locus.

**Supplementary Table 2b**: Annotations for variants within GWAS significant Pleiotropic Genomic Risk Loci for Discordant Variants

*Note:* uniqID : Unique ID of SNPs consisting of chr:position:allele1:allele2 where alleles are alphabetically ordered; rsID : rsID based on dbSNP build 146; chr : chromosome; pos : position on hg19; effect_allele : Effect/risk allele if it is provided in the input GWAS summary statistics file. If not, this is the alternative (minor) allele in 1000G; non_effect_allele : Non-effect/non-risk allele if it is provided in the input GWAS summary statistics file. If not, this is the reference (major) allele in 1000G; MAF : Minor allele frequency computed based on 1000G; gwasP : P-value provided in the input GWAS summary statistics file. Non-GWAS tagged SNPs (which do not exist in input file but are extracted from the reference panel) have "NA" instead; or : Odds ratio provided in the input GWAS summary statistics file if available. Non-GWAS tagged SNPs (which do not exist in input file but are extracted from the reference panel) have "NA" instead; beta : Beta provided in the input GWAS summary statistics file if available. Non-GWAS tagged SNPs (which do not exist in input file but are extracted from the reference panel) have "NA" instead; se : Standard error provided in the input GWAS summary statistics file if available. Non-GWAS tagged SNPs (which do not exist in input file but are extracted from the reference panel) have "NA" instead; r2 : The maximum r2 of the SNP with one of the independent significant SNPs; IndSigSNP : rsID of the independent significant SNP which has the maximum r2 with the SNP; Genomic locus : Index of the genomic risk loci matching with "Genomic risk loci" table; Novel : Whether the index SNP/locus is novel to the pleiotropic analysis, and if the variant/locus has been validated in recent education attainment GWAS (information from Supplementary Table 4); nearestGene : The nearest Gene of the SNP based on ANNOVAR annotations. Note that ANNOVAR annotates "consequence" function by prioritizing the most deleterious annotation for SNPs which are locating a genomic region where multiple genes are obverlapped. Genes are ecoded in symbol, if it is available otherwise; Ensembl ID. Genes include all transcripts from Ensembl gene build 85 including non-protein coding genes and RNAs; dist : Distance to the nearest gene. SNPs which are locating in the gene body or 1kb up- or down-stream of TSS or TES have 0; func : Functional consequence of the SNP on the gene obtained from ANNOVAR. For exonic SNPs, detailed annotation (e.g. non-synonymous, stop gain and so on) is available in the ANNOVAR table (annov.txt); CADD : CADD score which is computed based on 63 annotations. The higher the score, the more deleterious the SNP is. 12.37 is the suggested threshold by Kicher et al (2014); RDB : RegulomeDB score which is a categorical score (from 1a to 7). 1a is the highest score for SNPs with the most biological evidence to be a regulatory element; minChrState : The minimum 15-core chromatin state across 127 tissue/cell type; commonChrState : The most common 15-core chromatin state across 127 tissue/cell types; posMapFilt : Whether the SNP was used for eQTL mapping or not. 1 is used, otherwise 0. When eqtl mapping is not performed, all SNPs have 0.

**Supplementary Table 2c:** GWAS significant variants within r^2^ = 0.6 of Discordant sentinel variants

*Note:* No : Index of independent significant SNPs; Genomic Locus : Index of assigned genomic locus matched with "Genomic risk loci" table. Multiple independent lead SNPs can be assigned to the same genomic locus; uniqID : Unique ID of SNPs consisting of chr:position:allele1:allele2 where alleles are alphabetically ordered; rsID : rsID based on dbSNP build 146; chr : chromosome; pos : position on hg19; P-value : P-value (from the input file); nSNPs : The number of SNPs in LD with the lead SNP given r2, including non-GWAS-tagged SNPs (which are extracted from 1000G); nGWASSNPs : The number of GWAS-tagged SNPs in LD with the lead SNP given r2. This is a subset of "nSNPs".

**Supplementary Table 2d**: GWAS significant Pleiotropic Discordant sentinel variants within a given genomic locus

*Note:* No : Index of lead SNPs; Genomic Locus : Index of assigned genomic locus matched with "Genomic risk loci" table. Multiple lead SNPs can be assigned to the same genomic locus; uniqID : Unique ID of SNPs consisting of chr:position:allele1:allele2 where alleles are alphabetically ordered; rsID : rsID based on dbSNP build 146; chr : chromosome; pos : position on hg19; P-value : P-value (from the input file); nIndSigSNPs : Number of independent significant SNPs which are in LD with the lead SNP at r2 0.1; IndSigSNPs : rsID of independent significant SNPs which are in LD with the lead SNP at r2 0.1.

**Supplementary Table 3a**: GWAS significant Pleiotropic Genomic Risk Loci for “Dual” Variants

*Note:* Genomic locus : Index of genomic rick loci; uniqID : Unique ID of SNPs consisting of chr:position:allele1:allele2 where alleles are alphabetically ordered; rsID : rsID of the top lead SNP based on dbSNP build 146; chr : chromosome of top lead SNP; pos : position of top lead SNP on hg19; P-value : P-value of top lead SNP (from the input file); start : Start position of the locus; end : End position of the locus; nSNPs : The number of unique candidate SNPs in the genomic locus, including non-GWAS-tagged SNPs (which are available in the user selected reference panel). Candidate SNPs are all SNPs that are in LD (give user-defined r2) with any of independent significant SNPs and either have a P-value below the user defined threshold or are only available in 1000G; nGWASSNPs : The number of unique GWAS-tagged candidate SNPs in the genomic locus which is available in the GWAS summary statistics input file. This is a subset of "nSNPs"; nIndSigSNPs : The number of the independent (at user defined r2) significant SNPs in the genomic locus; IndSigSNPs : rsID of the independent significant SNPs in the genomic locus; nLeadSNPs : The number of lead SNPs in the genomic locus. Lead SNPs are subset of independent significant SNPs at r2 0.1; LeadSNPs : rsID of lead SNPs in the genomic locus.

**Supplementary Table 3b**: Annotations for variants within GWAS significant Pleiotropic Genomic Risk Loci for “Dual” Variants

*Note:* uniqID : Unique ID of SNPs consisting of chr:position:allele1:allele2 where alleles are alphabetically ordered; rsID : rsID based on dbSNP build 146; chr : chromosome; pos : position on hg19; effect_allele : Effect/risk allele if it is provided in the input GWAS summary statistics file. If not, this is the alternative (minor) allele in 1000G; non_effect_allele : Non-effect/non-risk allele if it is provided in the input GWAS summary statistics file. If not, this is the reference (major) allele in 1000G; MAF : Minor allele frequency computed based on 1000G; gwasP : P-value provided in the input GWAS summary statistics file. Non-GWAS tagged SNPs (which do not exist in input file but are extracted from the reference panel) have "NA" instead; or : Odds ratio provided in the input GWAS summary statistics file if available. Non-GWAS tagged SNPs (which do not exist in input file but are extracted from the reference panel) have "NA" instead; beta : Beta provided in the input GWAS summary statistics file if available. Non-GWAS tagged SNPs (which do not exist in input file but are extracted from the reference panel) have "NA" instead; se : Standard error provided in the input GWAS summary statistics file if available. Non-GWAS tagged SNPs (which do not exist in input file but are extracted from the reference panel) have "NA" instead; r2 : The maximum r2 of the SNP with one of the independent significant SNPs; IndSigSNP : rsID of the independent significant SNP which has the maximum r2 with the SNP; Genomic locus : Index of the genomic risk loci matching with "Genomic risk loci" table; Novel : Whether the index SNP/locus is novel to the pleiotropic analysis, and if the variant/locus has been validated in recent education attainment GWAS (information from Supplementary Table 4); nearestGene : The nearest Gene of the SNP based on ANNOVAR annotations. Note that ANNOVAR annotates "consequence" function by prioritizing the most deleterious annotation for SNPs which are locating a genomic region where multiple genes are obverlapped. Genes are ecoded in symbol, if it is available otherwise; Ensembl ID. Genes include all transcripts from Ensembl gene build 85 including non-protein coding genes and RNAs; dist : Distance to the nearest gene. SNPs which are locating in the gene body or 1kb up- or down-stream of TSS or TES have 0; func : Functional consequence of the SNP on the gene obtained from ANNOVAR. For exonic SNPs, detailed annotation (e.g. non-synonymous, stop gain and so on) is available in the ANNOVAR table (annov.txt); CADD : CADD score which is computed based on 63 annotations. The higher the score, the more deleterious the SNP is. 12.37 is the suggested threshold by Kicher et al (2014); RDB : RegulomeDB score which is a categorical score (from 1a to 7). 1a is the highest score for SNPs with the most biological evidence to be a regulatory element; minChrState : The minimum 15-core chromatin state across 127 tissue/cell type; commonChrState : The most common 15-core chromatin state across 127 tissue/cell types; posMapFilt : Whether the SNP was used for eQTL mapping or not. 1 is used, otherwise 0. When eqtl mapping is not performed, all SNPs have 0.

**Supplementary Table 3c:** GWAS significant variants within r^2^ = 0.6 of “Dual” sentinel variants

*Note:* No : Index of independent significant SNPs; Genomic Locus : Index of assigned genomic locus matched with "Genomic risk loci" table. Multiple independent lead SNPs can be assigned to the same genomic locus; uniqID : Unique ID of SNPs consisting of chr:position:allele1:allele2 where alleles are alphabetically ordered; rsID : rsID based on dbSNP build 146; chr : chromosome; pos : position on hg19; P-value : P-value (from the input file); nSNPs : The number of SNPs in LD with the lead SNP given r2, including non-GWAS-tagged SNPs (which are extracted from 1000G); nGWASSNPs : The number of GWAS-tagged SNPs in LD with the lead SNP given r2. This is a subset of "nSNPs".

**Supplementary Table 3d**: GWAS significant Pleiotropic “Dual” sentinel variants within a given genomic locus

*Note:* No : Index of lead SNPs; Genomic Locus : Index of assigned genomic locus matched with "Genomic risk loci" table. Multiple lead SNPs can be assigned to the same genomic locus; uniqID : Unique ID of SNPs consisting of chr:position:allele1:allele2 where alleles are alphabetically ordered; rsID : rsID based on dbSNP build 146; chr : chromosome; pos : position on hg19; P-value : P-value (from the input file); nIndSigSNPs : Number of independent significant SNPs which are in LD with the lead SNP at r2 0.1; IndSigSNPs : rsID of independent significant SNPs which are in LD with the lead SNP at r2 0.1.

**Supplementary Table 4**: Novel Loci from PLEIO pleiotropic meta-analysis and Validation with recent Education Attainment GWAS

*Note:* Genomic Coordintes/Locus Information: lociID: LocusID in the format chr:start:stop; chrom : chromosome; start : start base pair coordinate of locus; end : end base pair coordinate of locus. Input GWAS: PGC_SCZ: more recent schizophrenia GWAS accessed from PGC’s website, Edu_Lee: 2019 data freeze of SSGAC GWAS accessed from SSGAC’s website, LAM_CTP: cognitive data access via available via COGENT data links available from Lam et al., 2022 Nat Comm manuscript; Education (Validation): Most recent 2023 Education GWAS significant SNPs accessed from SSGAC’s website; further details in Data Availability section; Status: Novel: loci that are not previously reported on in the input GWAS; Novel replicated: loci that are note previously reported on in the input GWAS, but validated in most recent Education Attainment GWAS; Pleiotropy Meta-Analysis: Concordant: output of pleiotropic meta-analysis of concordant variant set; Discordant: output of pleiotropic meta-analysis of discordant variant set; Dual: output of pleiotropic meta-analysis of dual variant set; index variant: index variant within significant genomic locus; pval: p-value of index variant.

**Supplementary Table 5a**: MAGMA Gene Set Analysis for Concordant Variant Set

*Note:* FULL_NAME: name of gene set; Total_NumberGenes: total number of genes within each gene set;	NGENES: number of genes included from MAGMA gene-based genome-wide analysis; BETA: effect size of MAGMA gene set analysis; BETA_STD: standard deviation of effect size; SE: standard error of gene set analysis; P: uncorrected p-value; P_bonferroni_corr: Bonferroni corrected p-values; P_sidak_corr: Sidak corrected p-values; P_holm-sidak_corr: Holm-Sidak corrected p-values; P_fdr_bh_corr: Benjamini-Hochberg FDR corrected p-values; P_holm_corr: Holm corrected p-values; CommonGenes: gene included within MAGMA gene set analysis results; CommonGenes_Pvalues: MAGMA gene-based p-values

**Supplementary Table 5b**: MAGMA Gene Set Analysis for Discordant Variant Set

*Note:* FULL_NAME: name of gene set; Total_NumberGenes: total number of genes within each gene set;	NGENES: number of genes included from MAGMA gene-based genome-wide analysis; BETA: effect size of MAGMA gene set analysis; BETA_STD: standard deviation of effect size; SE: standard error of gene set analysis; P: uncorrected p-value; P_bonferroni_corr: Bonferroni corrected p-values; P_sidak_corr: Sidak corrected p-values; P_holm-sidak_corr: Holm-Sidak corrected p-values; P_fdr_bh_corr: Benjamini-Hochberg FDR corrected p-values; P_holm_corr: Holm corrected p-values; CommonGenes: gene included within MAGMA gene set analysis results; CommonGenes_Pvalues: MAGMA gene-based p-values

**Supplementary Table 5c**: MAGMA Gene Set Analysis for Dual Variant Set

*Note:* FULL_NAME: name of gene set; Total_NumberGenes: total number of genes within each gene set;	NGENES: number of genes included from MAGMA gene-based genome-wide analysis; BETA: effect size of MAGMA gene set analysis; BETA_STD: standard deviation of effect size; SE: standard error of gene set analysis; P: uncorrected p-value; P_bonferroni_corr: Bonferroni corrected p-values; P_sidak_corr: Sidak corrected p-values; P_holm-sidak_corr: Holm-Sidak corrected p-values; P_fdr_bh_corr: Benjamini-Hochberg FDR corrected p-values; P_holm_corr: Holm corrected p-values; CommonGenes: gene included within MAGMA gene set analysis results; CommonGenes_Pvalues: MAGMA gene-based p-values

### Supplementary Figures

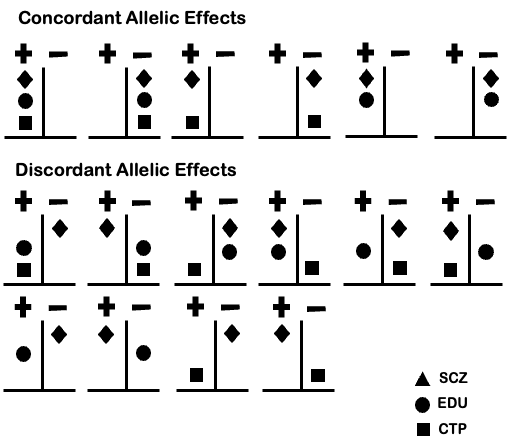

**Supplementary Figure 1**. Concordant and Discordant Effect Allele Sets

Top panel: Possible allelic configurations that constitute the Concordant allele variant set. Bottom panel: Possible allele configurations that constitute the Discordant allele variant set.
